# Issues of Random Sampling with Rapid Antigen Tests for COVID-19 Diagnosis: A Special Reference to Kalmunai RDHS Division

**DOI:** 10.1101/2021.01.11.21249636

**Authors:** AM Razmy, SM Junaideen

## Abstract

Random sampling from the community and performing the rapid antigen test has become a debatable issue during this COVID-19 pandemic. This writing analyzes the concerns using the data from Kalmunai RDHS Division, Sri Lanka, related to COVID-19 and shows the issues of random sampling in a community wherein the incident rate is small. Therefore, suitable sampling protocol is to be developed for performing rapid antigen test for COVID-19 diagnosis.

## 1. Introduction

Random sampling for diagnosing COVID-19 through rapid antigen test (RAT) is being carried out in different parts of the world. While the reliability of the test outcome is being widely debated, this writing attempts to elaborate the issues with random sampling in the community to detect COVID-19 infection through RAT. Mainly due to the specificity of testing kit, the reliability of the outcome is questioned. This issue creates complications in the community and health workers face numerous unwanted issues. The COVID-19 data available for Regional Directorate of Health Services (RDHS), Kalmunai, Sri Lanka, has been used for elaborating the associated risks.

## 2. Methodology

### 2.1 Data

The COVID-19 data available for RDHS, Kalmunai, Sri Lanka, up to 10.00 am on 10.01.2021, is given in Table 1. This RDHS division has the population of 443,577 and it is located in the eastern part of the Sri Lanka. The total number of diagnosis test carried out is 22,334 in the 13 Medical Officer of Health (MOH) sub divisions and these tests includes either PCR or RAT. The last two columns of this table give the number of incidents per 10,000 individuals and the number of tests done per 1,000 individuals for each MOH sub divisions.

**Table 1.** COVID-19 data for the Kalmunai RDHS Division as at 10.00 am on 10.01.2021.

| **MOH** | **Population** | **Positive Cases** | **TEST (PCR & RAT)** | **Incident per 10,000** | **Test per 1,000** |
| --- | --- | --- | --- | --- | --- |
| AKARAIPATHU | 43392 | 310 | 4983 | 71.4 | 114.8 |
| KALMUNAI(S) | 49488 | 229 | 3936 | 46.3 | 79.5 |
| POTTUVIL | 38564 | 77 | 1272 | 20.0 | 33.0 |
| SAINTHAMARUTHU | 28207 | 56 | 938 | 19.9 | 33.3 |
| ADDALAICHCHNAI | 46495 | 88 | 2156 | 18.9 | 46.4 |
| IRRAKAMAM | 15935 | 24 | 593 | 15.1 | 37.2 |
| ALYADIVEMPU | 24880 | 36 | 2191 | 14.5 | 88.1 |
| KARAITHIVU | 18656 | 21 | 1183 | 11.3 | 63.4 |
| NAVITHNAVELY | 20748 | 16 | 549 | 7.7 | 26.5 |
| NINTAVUR | 29205 | 18 | 1346 | 6.2 | 46.1 |
| THIRUKOIL | 28004 | 15 | 840 | 5.4 | 30.0 |
| KALMUNIA(N) | 33015 | 17 | 1322 | 5.1 | 40.0 |
| SAMMANTHURAI | 66988 | 27 | 1025 | 4.0 | 15.3 |
| **TOTAL** | **443577** | **934** | **22334** | **21.1** | **50.3** |

*(Source: Kalmunai RDHS division)*

### 2.2 Sensitivity and Specificity of the RAT Kit

There are many studies on the sensitivity [the rate of detecting infections correctly i.e., P (+/really positive)] and specificity [the rate of detecting non infections correctly i.e., P (-/really negative)] of the RAT. When the RAT is used on people who were positive for COVID-19 in a standard PCR test, Abbott’s antigen assay correctly spotted the virus in 95–100% of cases if the samples were collected within a week of the onset of symptoms. But that proportion dropped to 75% if samples were taken more than a week after people first showed symptoms. The sensitivity of the other RAT used in the United States is between 84% and 98% if a person is tested in the week after showing symptoms [1]. European centre of disease prevention and control (ECDC) has performed a meta-analysis of the clinical performance of commercial four RATs and retrieved additional results of clinical evaluation studies of nine RATs from eight companies. This meta-analysis resulted the sensitivities and specificities against RT-PCR tests and ranged between 29% (95% CI 15.7-42.3) and 93.9% (95% CI 86.5-97.4) for test sensitivity and between 80.2% (95% CI 71.1-86.7) and 100% (95% CI 98.8-100) for test specificity [2]. The differences in performance noted between the tests and between the studies can be partially explained by different populations and time of testing (proportion of persons that were tested early versus late in the course of the disease), and may also be affected by different RT-PCR assays used as gold-standard comparators, extraction methods or type of samples [3]. The WHO [4], Health Canada [5] and the US Centers for Disease Control and Prevention have recently issued guidelines for the use of RATs [6]. The WHO recommends RATs that meet the minimum performance requirements of ≥80% sensitivity and ≥97% specificity [4], while ECDC suggests aiming to use tests with a performance closer to RT-PCR, i.e., ≥90% sensitivity and ≥97% specificity [3]. Since the WHO South-East Asia Regional Office and the WHO Sri Lanka supplied RAT kits to Sri Lanka [7], it is reasonable to assume the sensitivity and specificity of the RAT used in the Kalmunai RDHS division are 80% and 97% respectively.

## 3. Analysis

Figure 1 shows the COVID-19 incidents per 10,000 and the number of tests carried out per 1,000 for each MOH sub divisions in the Kalmunai RDHS division. In general, the incident rate has increased with the test rate in the population with some exceptions such as for the Alayadivempu, Karaithivu MOH sub divisions. This can be further investigated by the scatter diagram given in figure 2.

**Figure 1.**
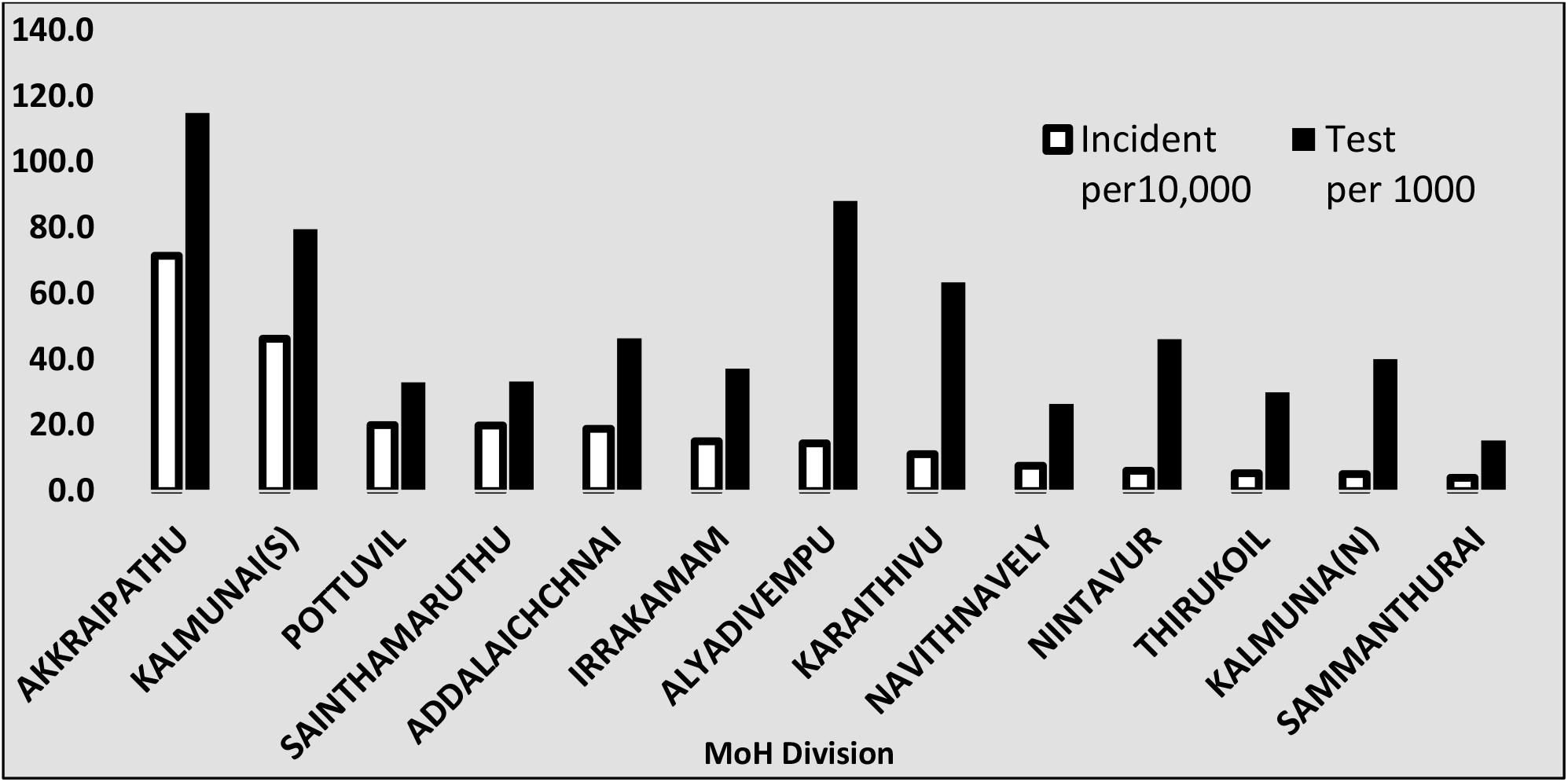
Covid-19 incidents and test rate.

**Figure 1.**
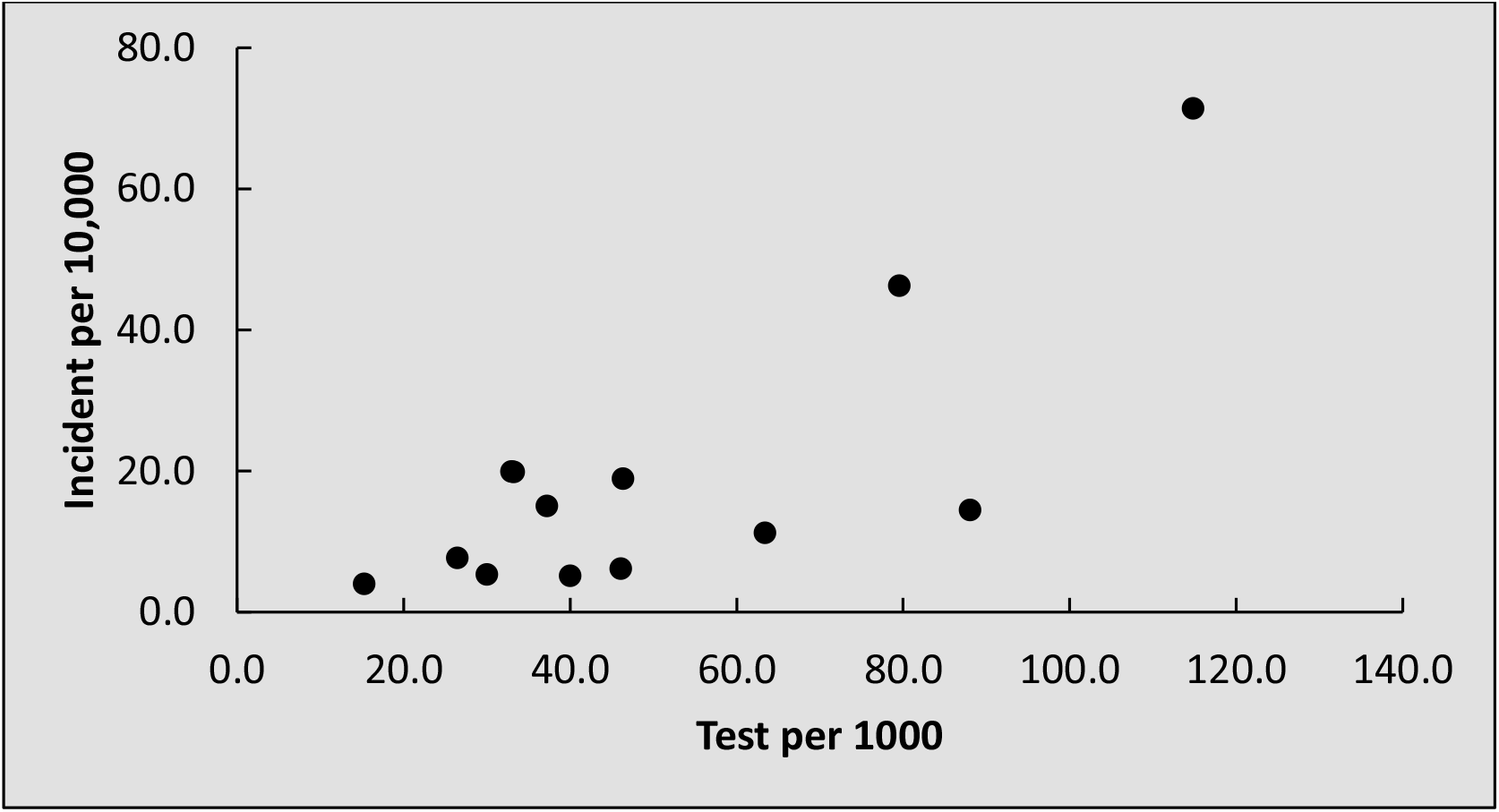
Scatter diagram to show the relationship between incidents and the test rate.

If one assumes these tests are done at random, an enquiry strikes that the incident rate has increased with the increasing test rates. This paper is answering to this issue when using the RATs which has the sensitivity of 80% [Probability (+/really positive) = *P*(+/*C*) = 0.80] and the specificity of 97% [Probability (-/really negative) = *P*(−/*C*′) = 0.97], used in the Kalmunai RDHS division. The total number of positive cases identified in the Kalmunai RDHS division is 934. Therefore, the incident rate can be considered as 0.0418 [The Probability (P) for the COVID-19 in the community = *P*(*C*) = 0.0418] assuming the tests were done at random. Considering these statistics, then the probability that an individual truly has COVID-19 given that a RAT has a positive result [*P*(*C*/+)] is

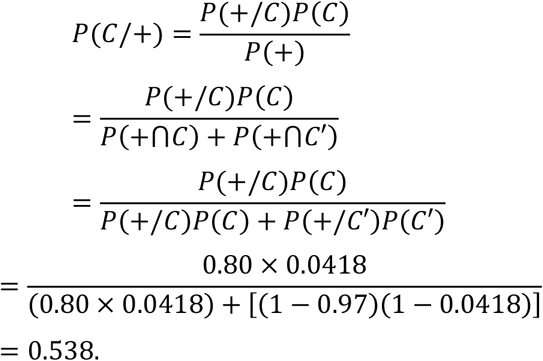

## 4. Results and Discussion

The analysis in section 3 says, in a community with 0.0418 incident rate, if the RATs are done randomly, the chance of that diagnosed person having COVID-19 in reality is only 53.8 % and 46.2% of the positive diagnosis are done wrongly. It has to be noted that the incident rate was estimated as 0.0418 assuming all the tests were done randomly but in reality, it is not. Purposive samples were also there in the total of 22,334 samples. Therefore, this estimated incident rate should be less than the value of 0.0418, which will increase the false positives further. This result can be enlightened by the figure 3 based on the statistics available. In a community with 10,000 individuals, there will be a large number of individuals (≈ 9582) without COVID-19 against the ≈ 418 COVID-19 patients. If all 10,000 individuals are tested with RAT, with the said sensitivity, ≈ 335 of the 418 COVID-19 patients will be diagnosed for COVID-19. With the said specificity, ≈ 228 individuals of the 9,582 non COVID-19 individuals will diagnosed for COVID-19 wrongly. This proves that when the incident rate is small in the community and the test kits are used at randomly, large number of false positives cases can be observed.

**Figure 3.**
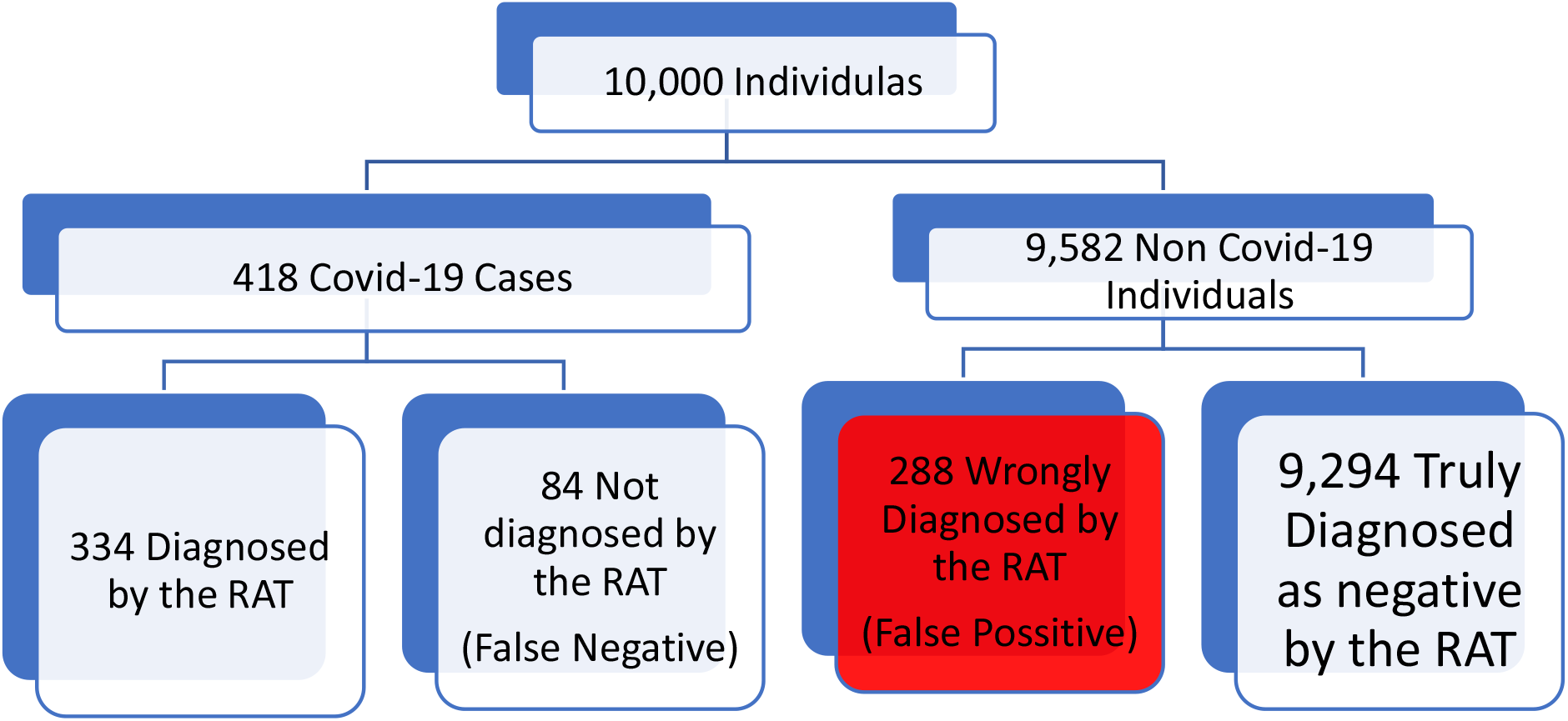
Explanation for the results.

## 5. Conclusion

There are large number of non-COVID-19 cases in the population (95.8%). Since the test has 97 % specificity, 3 % of the non-COVID-19 cases are falsely identified as case. This issue drastically reduces the reliability of the test to 53.8 % when one gets positive diagnosis. This reliability will go further down since the estimated incident rate was based on the assumption of random sampling which is not true in every case. Therefore, a suitable protocol is required for the sampling the mechanism when performing RATs in the community. However, the figure 1 can’t be an absolute example for question arisen that more tests-more positive since all sample were not obtained at random and PCR tests were also used for the diagnosis.

## Data Availability

This study is based on the published secondary data and therefore it is exempted. Data available in the official website of the Ministry of health

http://www.health.gov.lk/moh_final/english/others.php?pid=98

